# Evaluating the Human Safety Net: Observational study of Physician Responses to Unsafe AI Recommendations in high-fidelity Simulation

**DOI:** 10.1101/2023.10.03.23296437

**Authors:** Paul Festor, Myura Nagendran, Anthony C. Gordon, A. Aldo Faisal, Matthieu Komorowski

**Author notes:** Equal contribution (Paul Festor, Myura Nagendran). These authors jointly supervised this work (A. Aldo Faisal, Matthieu Komorowski).

## Abstract

In the context of Artificial Intelligence (AI)-driven decision support systems for high-stakes environments, particularly in healthcare, ensuring the safety of human-AI interactions is paramount, given the potential risks associated with erroneous AI outputs. To address this, we conducted a prospective observational study involving 38 intensivists in a simulated medical setting.

Physicians wore eye-tracking glasses and received AI-generated treatment recommendations, including unsafe ones. Most clinicians promptly rejected unsafe AI recommendations, with many seeking senior assistance. Intriguingly, physicians paid increased attention to unsafe AI recommendations, as indicated by eye-tracking data. However, they did not rely on traditional clinical sources for validation post-AI interaction, suggesting limited “debugging.”

Our study emphasises the importance of human oversight in critical domains and highlights the value of eye-tracking in evaluating human-AI dynamics. Additionally, we observed human-human interactions, where an experimenter played the role of a bedside nurse, influencing a few physicians to accept unsafe AI recommendations. This underscores the complexity of trying to predict behavioural dynamics between humans and AI in high-stakes settings.

## INTRODUCTION

Artificial Intelligence (AI) driven systems are set to take an increasingly prominent supportive role in decision-making including in high-stakes settings^1,2^. While the final decision remains in human hands, understanding how AI recommendations impact their user’s behaviour is crucial. In fact, recent work has highlighted differences in our perception of human and AI advice.^3,4^ In high-risk shared decision-making settings where even non-autonomous AI-decision-support tools have surprisingly high safety assurance requirements,^5^ understanding the human-AI dynamic is key to assessing overall safety.

In this paper, we look at live safety assessment of AI for decision support in healthcare because it combines many of the challenges that make AI safety assessment hard across the board. Whilst AI recommendation engines have shown promising results on retrospective data, the translation to the bedside has been slow due in part to concerns for patient safety.^6^ In situations where the optimal decision has historically been unclear, ^7–9^ and with technologies like reinforcement learning that aim at recommending decisions which surpass the standard of care, the safety-assessment challenge becomes even harder. One such example is cardiovascular management during sepsis where the optimal personalised doses of intravenous (IV) fluids and vasopressors remain unknown.^10,11^

Attempts have been made to improve the safety profile of AI-driven decision support in retrospective intensive care settings.^12–14^ Still, the necessity of prospective and higher fidelity evaluations involving clinical end users is clear from recent examples in other fields.^13^ For instance, an acute kidney injury alert system showing good performance on retrospective data was found to worsen outcomes when deployed in a real-world setting, illustrating the need for a careful transition between retrospective testing and prospective deployment of digital systems.^15^ Safely transitioning from “bytes to bedside” is a particularly complex challenge because of the dynamic interaction with human users who are prone to biases and can behave in unpredictable ways.^16–18^

In response to the growing emphasis on ecologically valid testing of AI systems,^19,20^ we run our behavioural experiment in a physical simulation suite, a tool which has historically been used as a widely accepted training tool for modelling high-fidelity situations and capturing patterns of human behaviour with simulation now forming a core part of medical training.^21^ This immersive approach enables physicians to respond to bedside stimuli more realistically, aligning their behaviours with actual clinical practice.^22^ Furthermore, this shift towards a more realistic setting aligns with the evolving regulatory landscape surrounding AI, which emphasises “human-centred AI” and the holistic evaluation of human-AI team performance.^23,24^ AI safety assessment is not a mere problem of computer science but also one of human-AI cooperation which should incorporate behavioural elements grounded in human perceptual and decision-making studies.

We use the example of cardiovascular management in sepsis to study the behaviour of physicians in response to AI recommendations. Here, we share the results of an observational study of human-AI interaction in a high-fidelity simulation suite focusing on the influence of safe and unsafe AI recommendations on treatment decisions. Using eye-tracking as a behavioural marker, we show evidence that the attention placed by clinicians on AI recommendations, as well as broader behavioural traits such as desire for senior advice, depends on AI advice safety. We also demonstrate that most unsafe AI recommendations would be appropriately rejected by the clinical team and recommend how clinical users of AI should be trained to further improve their robustness to hazardous AI recommendations.

## RESULTS

We conducted an observational human-AI interaction investigation within a high-fidelity simulation facility. Our primary aim was to assess clinicians’ ability to detect and appropriately reject unsafe AI recommendations or seek senior assistance. Participants engaged in simulated patient scenarios, prescribing fluid and vasopressor doses before and after receiving AI guidance, see Figure 1. The scenarios encompassed safe, unsafe, and “challenged” unsafe AI recommendations, with an experimenter acting as a bedside nurse trying to change the clinician’s decision in the latter case. Eye-tracking technology was used to monitor participants’ gaze patterns during simulations. Calibration and validation procedures ensured accurate gaze data collection. We divided the visual field into regions of interest (ROIs) to quantify attention patterns. Physicians were recruited from an ICU, and ethical approval was obtained. Data analysis was conducted using Python. The full study details, including the recruitment process and ethics approval, can be found in the Methods section.

**Figure 1:**
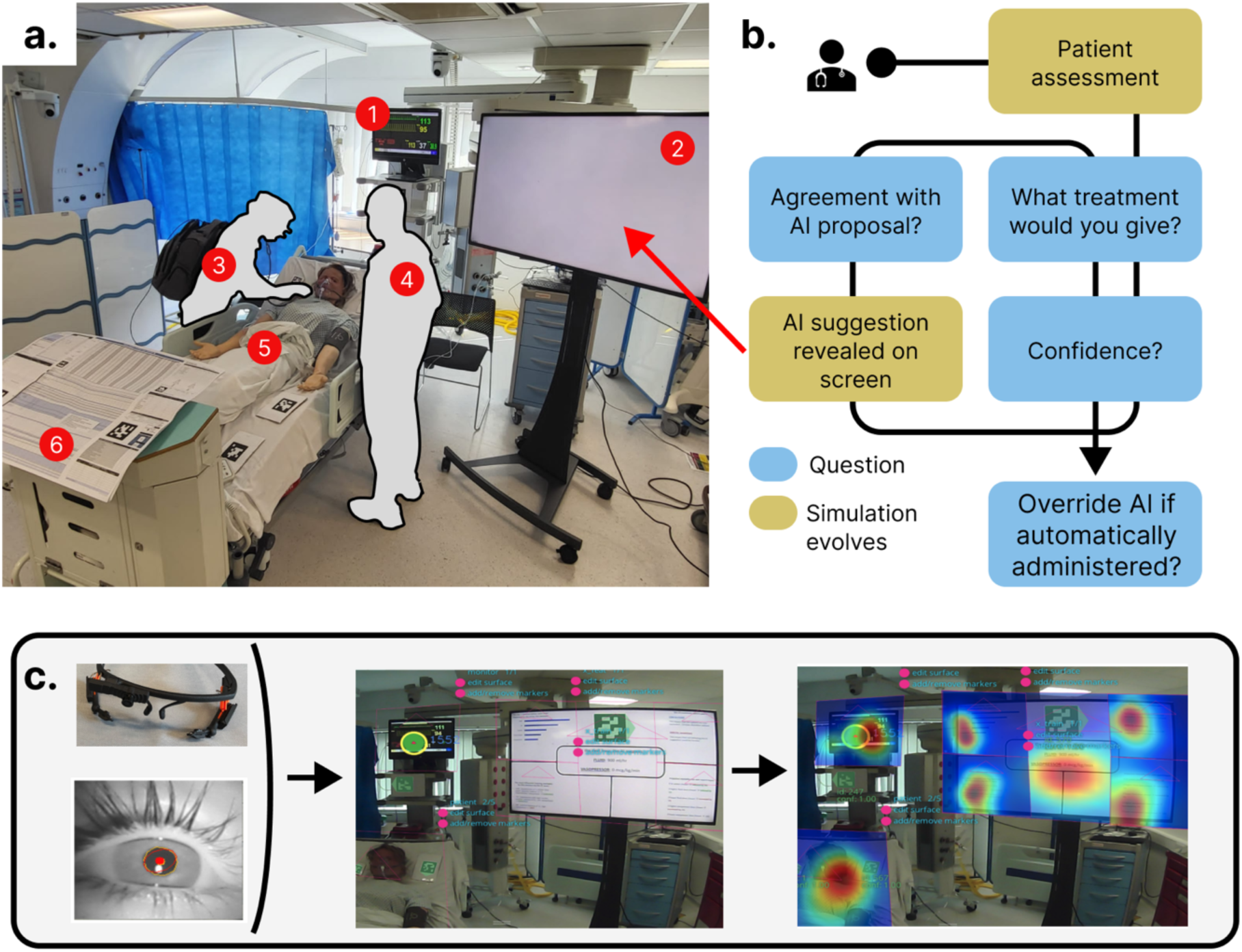
Experimental design. **A** Photo of the simulation suite with: (1) Bedside monitor (2) AI screen (3) Participant (4) Bedside nurse (5) Patient mannequin (6) Intensive care unit (ICU) bedside information chart. **B.** Experimental protocol diagram. **C.** Gaze-based attention extraction pipeline: eye-tracking glasses, pupil camera view, a recorded field of view with April tags (QR codes) and reconstructed data with fixation heatmaps on the different regions of interest (ROIs).

A total of 38 intensive care physicians took part in the experiment (Figure 2). This cohort comprised 25 men (66%) and 13 women (34%), proportions in line with the national population of intensivists.^25^ The balance between junior and senior physicians (with less or more than 5 years of experience respectively) was even and 21% of participants reported having been personally involved or having had experience, in AI research.

**Figure 2:**
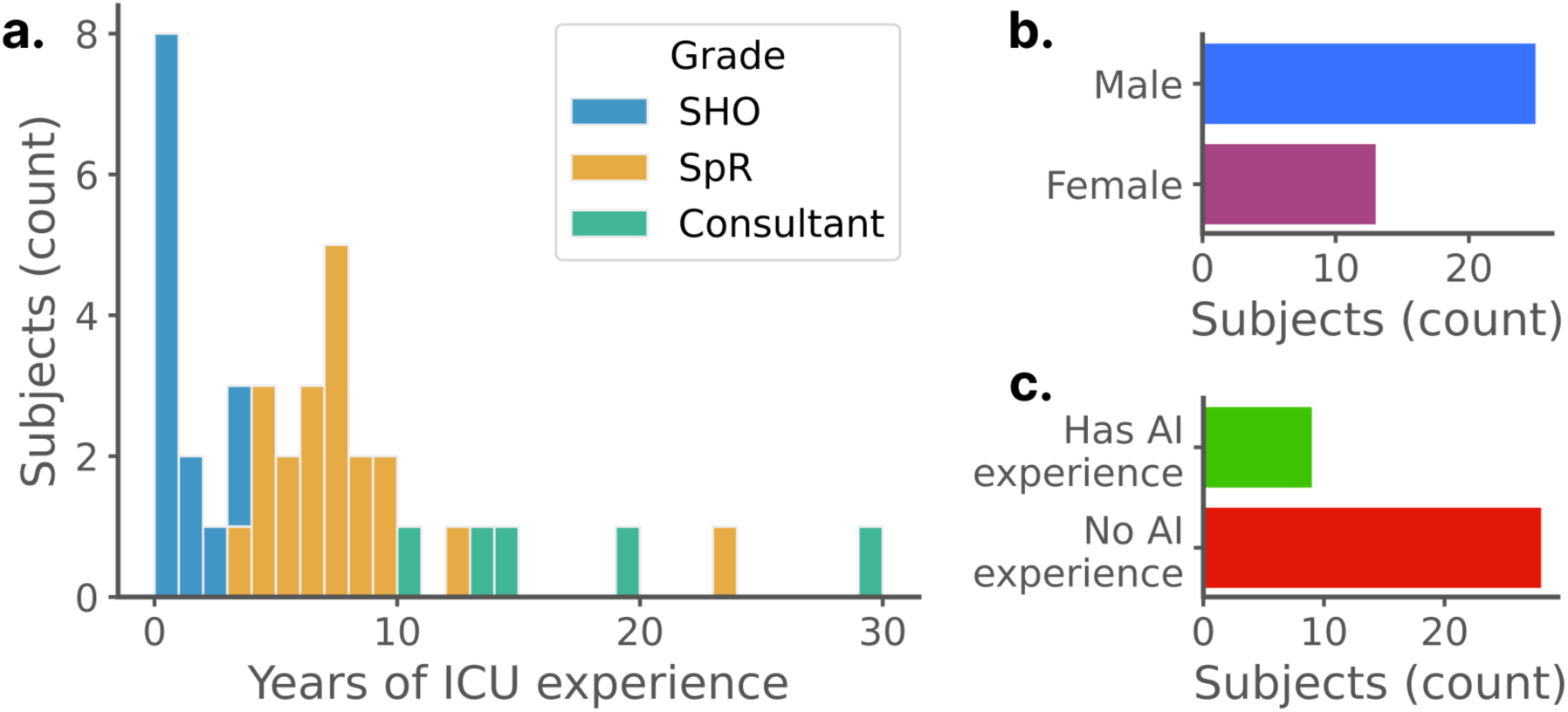
Recruited cohort demographics. **A** Distribution of intensive care experience. **B.** Gender distribution. **C.** Proportion of physicians who had ever been involved in a research project involving AI. This cohort covers the whole range of experience levels, is in line with national gender ratios and contains both people who have and have not worked with AI.

Each physician completed six (four safe, two unsafe) different patient scenarios leading to a total of 228 recorded trials. Of these trials, 76 featured an unsafe AI recommendation and 152 were safe ones. See Supplementary Appendix D for the full trial matrix.

In total, unsafe AI recommendations were stopped more often than safe AI recommendations (29% vs. 83%, p<0.0001). The proportion stopping unsafe AI recommendations rose to 92% (p=0.027) when including physicians who asked for a senior opinion, which would most likely lead to the unsafe AI recommendation being rejected (see Figure 3a). This analysis was further expanded by categorising physicians into junior (<5 years of intensive care unit (ICU) experience) and senior (≥5 years of ICU experience) practitioners. There was a non-significant trend for junior physicians to stop AI recommendations less often than senior physicians (79% vs. 83%, n.s.). Junior physicians asked more often for a second opinion than senior physicians (65% vs. 25%, p<0.0001), which led to more unsafe recommendations being stopped or escalated by juniors (94% by juniors against 91% by seniors, n.s.).

**Figure 3:**
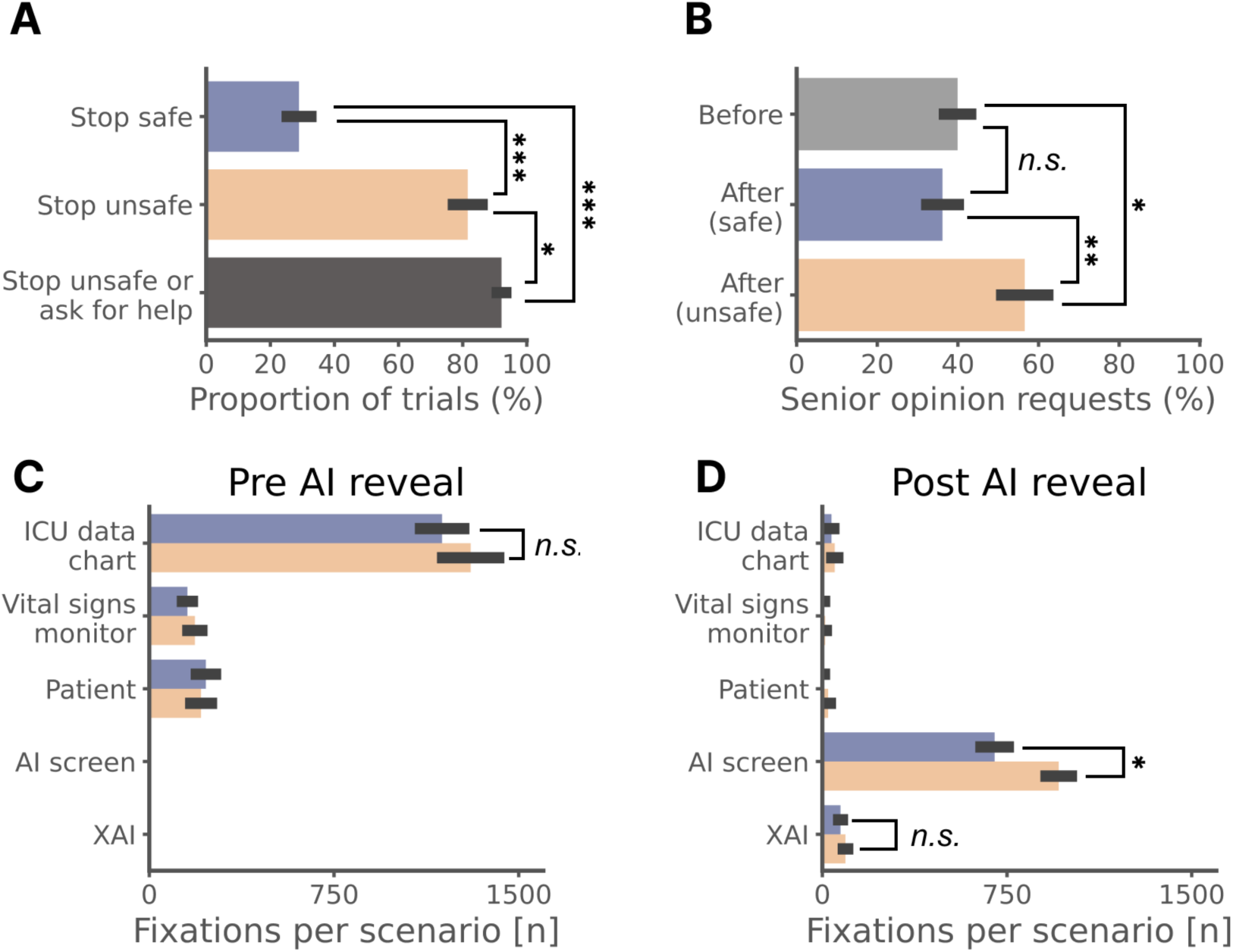
Impact of AI recommendation safety status on clinician decisions, and gaze fixations on each ROI. **A** Bar chart of the proportion of stopped safe recommendations, stopped unsafe recommendations and stopped or escalated unsafe recommendations. **B.** Proportions of requests for senior help before seeing any recommendation and after having seen a safe or an unsafe one. **C.** Number of gaze fixations on each ROI before revealing the AI recommendation (i.e. there can be no fixations on the AI) **D.** Number of gaze fixations on each ROI after revealing the AI recommendation (when clinicians have already evaluated the non-AI information sources and so would be expected to look at these much less). *** p<0.0001, ** p<0.005, * p<0.05, *n.s.* not significant.

Similarly, second-opinion requests rose from 40% before seeing any AI recommendation to 57% after seeing an unsafe AI recommendation (p=0.0056) but the reduction in requests after seeing a safe AI recommendation was not significant (figure 3b). Seeing an unsafe rather than a safe AI recommendation triggered more senior/second opinion requests (57% vs. 36%, p=0.0017). Seeing unsafe AI recommendations therefore significantly increased the proportion of requests for senior help.

As expected, prior to the AI recommendation being revealed, no significant difference in gaze fixations on regions of interest (ROIs) was observed between safe and unsafe scenarios regarding the three AI-independent regions (ICU data chart, vital signs monitor, and patient mannequin), see Figure 3c. Subsequent to the disclosure of the AI recommendation, there were more fixations on the AI screen in the unsafe scenarios (mean 960) versus safe scenarios (mean 700) (p=0.0015, see Figure 3d). Finally, the number of gaze fixations on the AI explanation ROI was not significantly different between safe and unsafe scenarios

The distributions of initial fluid and vasopressor dose prescriptions across participants in our six scenarios are shown in Figure 4. These results show wide variation in clinical practice, even when given the exact same information. Figure 4 suggests that the extent of the variation in prescribing might depend on case-specific features (e.g. in scenario 2, the patient had already had more fluids than in scenario 1 so physicians gave less fluid, or patient 5 had sepsis related to infected and leaky heart valves so physicians were more careful with both fluid and vasopressor), see Supplementary Appendix E for an extended discussion.

**Figure 4:**
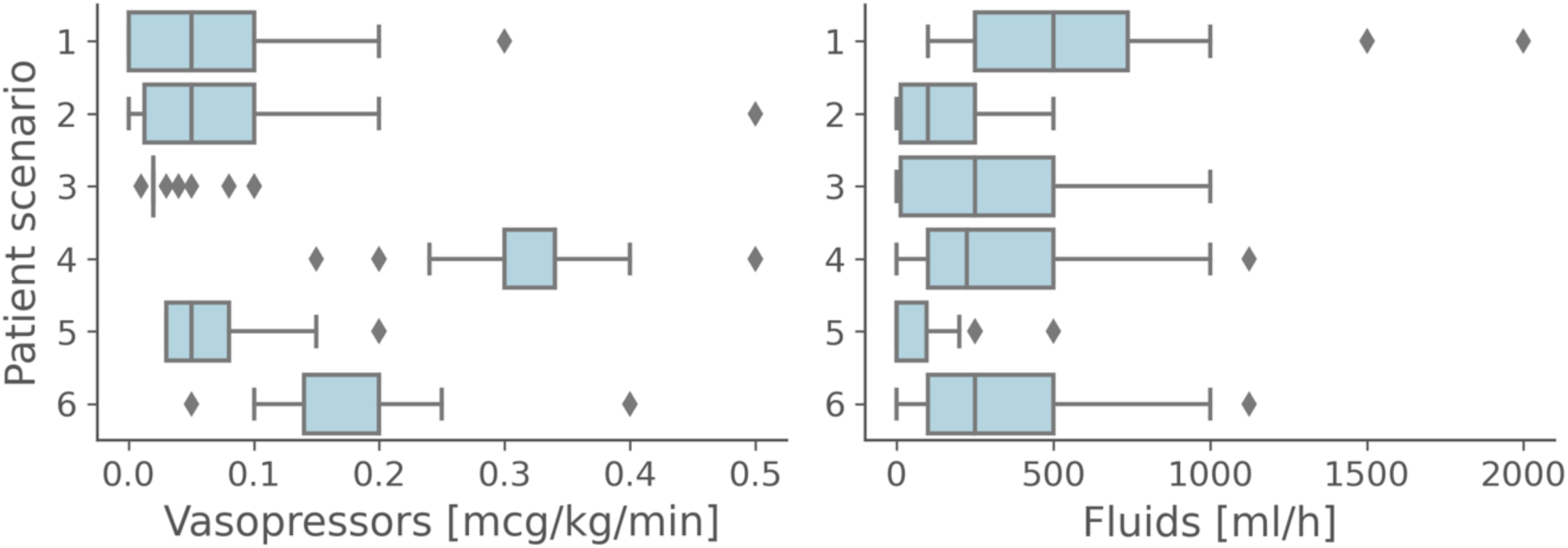
Clinical practice variability –. Distribution of initial (i.e. pre AI reveal) vasopressor (left) and fluid (right) prescriptions by physicians for each patient scenario.

We also investigated the extent to which AI recommendations influenced prescription decisions. Physicians changed their prescription (dose of fluids and/or vasopressors) in 46% (105/228) of trials after seeing what the AI suggested. Both safe and unsafe AI recommendations influenced human decisions to different extents: fluid doses shifted on average by 80 ml/h (and vasopressor doses by 0.01 mcg/kg/min) after a safe AI recommendation compared to 40 ml/h (and 0.08 mcg/kg/min) after an unsafe AI recommendation. Figure 5 shows the shift of distribution in vasopressor prescriptions before and after the AI recommendation was seen for two scenarios (split by whether the entire cohort is considered or only those physicians who did not ask for senior/second opinion). In scenarios (such as number two) where the unsafe AI recommendation was significantly influencing, this did not seem apparent when exclusively considering the physicians who did not request senior help (see Supplementary Appendix F for this plot over all scenarios).

**Figure 5:**
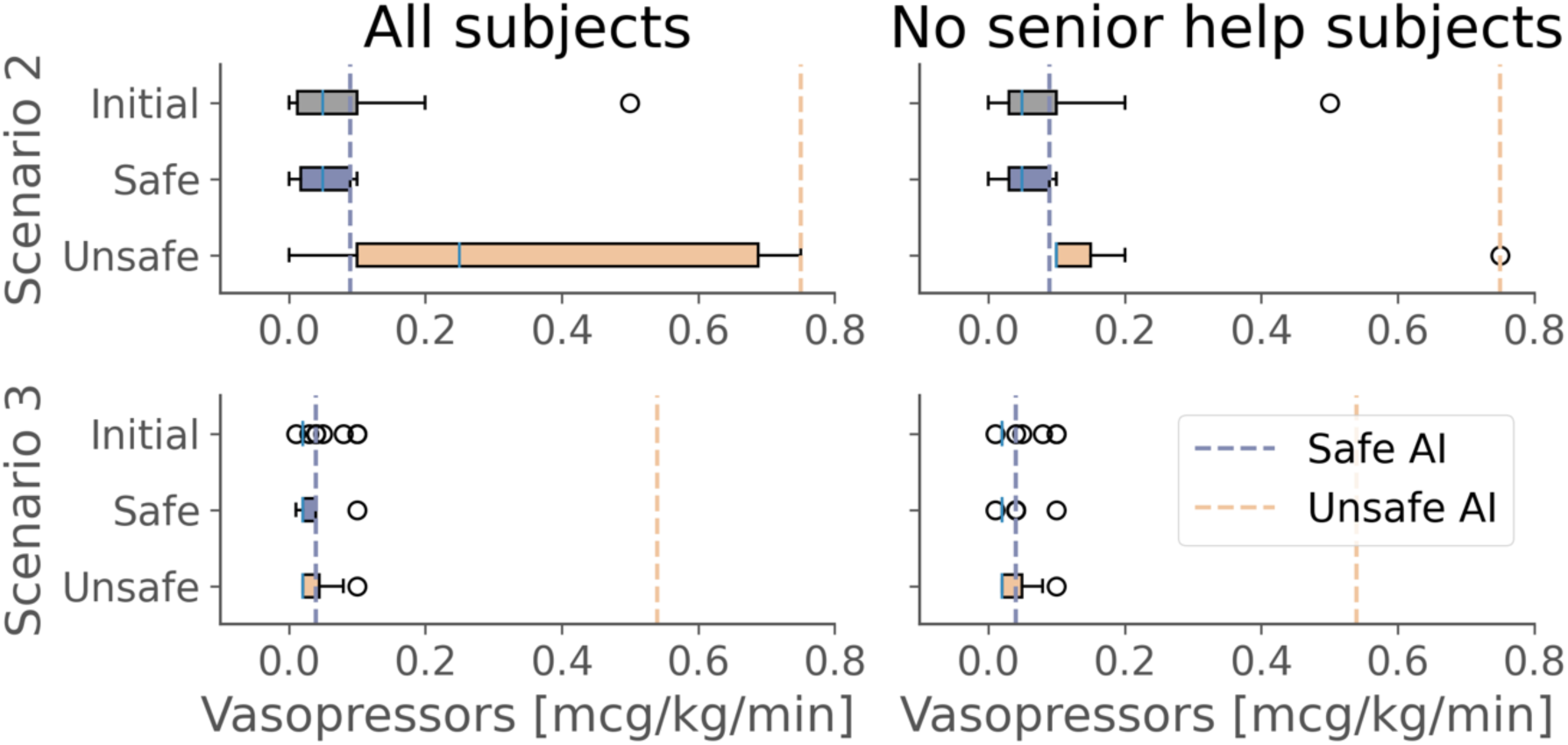
Shift analysis. Vasopressors dose distribution before and after having seen a safe or unsafe AI recommendation for two patient scenarios across all physicians (left) and only those who did not ask for senior/second review (right). Unsafe AI recommendations do not always influence the final decision. Physicians most influenced by unsafe AI recommendations tend to ask for a second opinion, while those who do not ask for help are less influenced by unsafe recommendations.

Finally, each physician had one of the two unsafe scenarios extended with a “challenge” section where the bedside nurse (a member of the experimental team) was given three attempts to change the physician’s mind on whether or not to stop the AI recommendation were it to be automatically implemented. In 95% of cases (36/38), the verbal input challenge from the bedside nurse did not sway the physician’s decision to accept or reject the automated application of the AI recommendation. However, two participants (both junior) were persuaded to change their minds from interrupting an unsafe recommendation to accepting it. Human-to-human interactions can therefore also play a role in the inadvertent adoption or appropriate rejection of unsafe recommendations.

## DISCUSSION

Our findings confirm that AI recommendations can influence clinician behaviour and thereby impact patient care. Unsafe AI recommendations, represented here as sudden under- or over-dosing, were frequently (but not entirely) detected and appropriately mitigated by the clinical team (by rejecting the AI recommendation). However, junior physicians more often deferred the decision to senior colleagues when they were unsure about the safety of an AI recommendation. This shows the importance of educating clinical teams who will interact with a new AI recommender system on the correct intended use of the system, including target patient population, indications, and limitations, as well as the importance of clinical context when integrating the AI recommendations into their practice.

This study reinforces the call for more interdisciplinary and realistic human-AI interaction studies on domain experts.^26,27^ Critically, our experimental design also allowed us to study human-human behavioural dynamics during an encounter with AI decision support. This is important as most clinical uses of AI-driven decision support tools will be in the context of multi-disciplinary teams where humans other than the final decision-maker can still positively or negatively influence the interaction between the final human decision-maker and the AI. This is why our study was run in a simulation suite: an environment that reproduces natural stimuli of bedside practice for clinicians without any risk to patients.

While eye-tracking is typically used in controlled environments,^28^ this study demonstrates the feasibility of using this behavioural phenotypic marker of attention in more realistic, less constrained, environments. Our findings indicate that physicians fixated more on unsafe than safe AI recommendations implying an appropriately higher level of allocated cognitive attention. However, we also observed that physicians did not rely more on AI explanations in the unsafe scenarios calling into question the use of explanations as a mitigation strategy for unsafe AI. Nor did physicians devote significantly more attention to looking back at the ‘traditional’ (non-AI based) clinical data after seeing an unsafe AI recommendation to understand why the recommendation might be unsafe (i.e. there was no outward evidence of a desire to ‘debug’ the unsafe AI recommendation).

The influence of AI recommendations on clinical judgement has already been studied in vignette-type experiments.^3^ This work goes one step closer to clinical deployment by studying these interactions in a high-fidelity simulation environment. This setup enabled the study of human-AI interaction with eye-tracking as well as the ability to investigate human-human interactions as they relate to AI. Most studies of clinical decision support system safety use medication error as the primary outcome measure and proxy for patient safety.^29^ Here, we look at systems that are not yet deployed in clinical practice, so measuring prescription error rate directly (and correlating that to ‘error’ without a gold standard) is challenging. Therefore, we took the problem from a different angle and aimed to estimate the ability of clinicians to spot unsafe treatment recommendations from an AI tool.

However, the limitations of the study should also be acknowledged. First, as raised by many physicians during the initial briefing, prescribing hourly fluid and vasopressor doses directly is unusual for intensive care physicians who typically indicate blood pressure (and other parameters) targets and let the bedside nurse titrate the actual doses within a reasonable range to reach the set targets. Similarly, the simulation limited the physician’s action space to one specific aspect of patient care, preventing action plans that might go beyond the defined possibilities. Moreover, making treatment decisions for the next hour is also less dynamic than real clinical practice (where for example the ability to examine a real patient and use advanced cardiac output monitors might add to the nuance of the overall clinical picture).

From a different perspective, one could challenge the definitions of safe and unsafe recommendations used in the scenarios by arguing that there is no ground truth in sepsis resuscitation and that they are therefore subjective. One might even go further and argue that strategies that under- or over-dose in specific patients (compared to the ‘average’) could be desirable in some cases. The scenarios used in this experiment were designed for the unsafe recommendations to be inappropriate to a majority of clinicians and validated by an independent panel of intensivists. The introduction of AI-driven decision support tools, particularly those using reinforcement learning aims to improve patient outcomes beyond the current standard of care.^9,30^ This means that such systems will give recommendations that differ from what the clinical team would ordinarily have done but potentially without explanation - a “mysterious oracle dilemma” where the AI oracle recommends actions that on average lead to better outcomes but might occasionally be suboptimal, and the users do not get context on the AI recommendation. It will therefore be essential for humans to exert critical thinking and assess how reasonable the AI recommendation is to filter potentially novel but superior calls by the AI from harmful recommendations.

As regulators push toward requiring clear intended purpose statements for software as medical devices,^31^ our high-fidelity eye-tracking based approach to evaluating an AI-driven decision support tool serves as a basis for promoting the generation of safety evidence. Furthermore, the recent rise in popularity of generative AI (most notably as large language models) highlights the safety concerns of hallucinatory outputs that might be acted upon in a clinical setting and bring harm to patients.^32^ It is likely that an AI system that shows overall superhuman performance in a given task will still show lower-quality performance in some specific cases.^33^ Solutions such as uncertainty-aware models or explainable AI might help users differentiate between well-informed recommendations and flawed calls.^34–36^ The human-AI interactions at the bedside, with a particular focus on high-pressure decision-making, would also help to accelerate the safe translation of AI-based decision support tools to the bedside.

## CONCLUSION

It is critical for clinician acceptance, regulatory compliance and real-world adoption that we evaluate cooperation between clinical experts and AI decision support tools in high-fidelity settings -in our case a simulated intensive care unit. This study demonstrates the influence of AI recommendations on clinical behaviour and suggests that the vast majority of unsafe AI recommendations are appropriately rejected by bedside clinicians. The findings on junior physicians occasionally accepting an unsafe AI recommendation and their general willingness to seek senior help when unsure should inform the intended use (i.e. some tools might need to only be used by junior clinicians if they have access to senior advice). Uncertainty awareness, novel forms of AI interpretability and a better understanding of human-human interactions (i.e. team decisions) in the context of AI-driven decision support will help not only with assuring safety from a regulatory perspective but also in fostering confidence and approval from physician end-users.

## METHODS

### Objective

We conducted an observational human-AI interaction study in a high-fidelity simulation facility. Our primary objective was to measure whether participants were able to detect, and correctly reject, unsafe recommendations from an AI tool and/or ask for senior help when appropriate. Secondary study objectives included: (i) quantifying the shift in fluid and vasopressor doses induced by seeing an AI recommendation, and (ii) determining whether or not gaze patterns varied differentially depending on the safety status of the AI recommendation.

### Experimental design

Participants (clinicians) were briefed on the experiment and completed a pre-experiment questionnaire recording their demographics and prior experience with AI (see Supplementary Appendix A for the full content of the briefing and questionnaire). Participants were told that they would conduct a review of several adult patients with sepsis within a simulation suite (Imperial College Simulation Centre) and that they would need to prescribe appropriate doses of fluid and vasopressor for each patient both before and after getting advice from an AI tool. Critically, physicians were told that the AI recommendation engine had been successfully validated in multiple retrospective settings but had not been prospectively evaluated. The simulation layout is shown in Figure 1a.

Each physician completed a total of six different patient scenarios, simulating a virtual “ward round”. Each scenario started with physicians entering the simulation suite and conducting their assessment of the patient as they saw fit. Data sources within the room included a standard paper ICU bedside data chart with observations and blood results, an ICU handover note including details of the patient’s presentation and medical history, a vital signs monitor and a physical patient mannequin (Simman 3G, Laerdal Medical, Stavanger, Norway) which could be examined (see Supplementary Appendix B for the details of each patient scenario). All patient scenarios were crafted by clinical experts from the authors team and to fit the needs of this simulation experiment, they do not come from real patients. A member of the research team played the role of the bedside ICU nurse who could only give standardised responses to any questions. Following their assessment of the patient, physicians were asked to recommend a dose rate for fluids (ml/hr) and vasopressors (noradrenaline, mcg/kg/min) for the coming hour (to match the format of AI recommendations). Physicians rated their confidence on a 1-10 scale and whether or not they would like support for their decision from a senior physician (or a second opinion if the physician was already senior themselves). They were then shown the AI recommendation, asked to what extent they agreed with the recommendation on a 5-point Likert scale (from completely disagree to completely agree), and then the initial dosing-related questions again (what dose they would prescribe, their confidence level, optional ask for senior help - see figure 1b).

Finally, physicians were asked whether or not they would stop the AI recommendation if it were to be automatically administered to the patient. This question was intended to nuance the agreement prompt and identify situations where a participant might disagree with an AI recommendation but not necessarily consider it a threat to patient safety. Participants were clearly introduced to the nuance between these two questions in the pre-experiment briefing.

The running of a single patient scenario from entry into the simulation suite to exit constituted one trial. Trials were categorised by the nature of the AI tool’s recommendation provided to the participant: safe, unsafe or “challenged” unsafe. In the latter, after the physician reported whether or not they would stop the AI recommendation if it were to be automatically administered, the bedside nurse was permitted three attempts (all following a standardised script) to verbally try to convince them to change their mind (see Supplementary Appendix C for the scripts). Each physician experienced four safe trials, one unsafe and one “challenged” unsafe in a pseudo-randomised order (see the trial matrix in Supplementary Appendix D). The first trial encountered by every physician was always in the safe condition to establish a baseline level of trust with the AI tool and let the physician familiarise themselves with the environment. The details of each patient scenario are presented in Supplementary Appendix B.

All AI recommendations were synthetically generated by the research team for the purpose of ensuring a standardised experimental format (i.e. they were not from a real AI system). The definition of unsafe recommendations was based on extreme under- or over-dosing of fluid and/or vasopressor as per previous work.^13^ All participating physicians were fully debriefed at the conclusion of the study on the synthetic nature of the AI recommendations so as not to bias their opinions of future interactions with AI-driven systems.

During each trial, all physician responses were recorded by a member of the research team sitting in a dead angle in the simulation suite. This data, along with questionnaire answers was reformatted and analysed in Python (code available online here: https://figshare.com/s/78c5ff5c6031f701c0d1)

### Eye-tracking for gaze recording

In this study, gaze was employed as an indicator of physicians’ attentional focus during simulations, with particular interest in whether this varied according to the safety of the AI recommendation. Pupil and first-person videos were recorded with non-invasive commercially available eye-tracking glasses (Pupil Core headset). The Pupil Labs software (Core, version 3.3) utilised both eye cameras to delineate the pupil and estimate the direction of gaze within the recorded field of view.

Prior to the experiment, a two-part 2D calibration procedure was conducted. The initial stage involved a static calibration using five screen markers on a laptop display (default Pupil Labs ‘screen marker’ calibration). Subsequently, a depth-based static exercise was performed, requiring participants to focus on nine screen markers sequentially (’natural features’ mode) displayed on a 60-inch TV screen, initially at 1 metre and then at 2 2-metre distance. A laptop (Lenovo Thinkpad) was connected to the eye-tracking glasses for the entire experiment. To allow for unrestricted movement in the suite, the glasses were connected via USB to a battery-powered laptop (Lenovo Thinkpad) worn by physicians in a lightweight backpack.

Because of variability in facial morphologies, 19/38 physicians passed the calibration exercises and had their gaze-based attention data collected. Physicians were instructed to point to where they were reading on the handover note at the start of each scenario as a final level of validation that the eye tracking was appropriately calibrated.

We defined four key regions of interest (ROIs) (Figure 1c): the paper ICU data chart, the vital signs monitor, the patient mannequin (Laerdal Simman 3G) and the AI display screen. Four further sub-regions were identified within the AI screen ROI corresponding to four types of explanation for the AI recommendation. April tags (simple QR codes) within the simulation suite (see Figure 1c) were used to identify ROIs in post-processing. As is common practice in eye-tracking literature ^37,38^, we used the number of gaze fixations per ROI—a fixation being the predominant eye movement occurring when the foveal region of the visual field is held stationary— as a proxy for participant attention.

### Participant recruitment and simulation facility

Recruitment of ICU physicians made use of both convenience sampling and targeted advertising to a local NHS trust (Imperial College Healthcare NHS Trust) Inclusion criteria were: (i) practising physician, (ii) has worked for two or more months in an adult ICU, (iii) currently works in ICU or has worked in ICU within the last 6 months. Physicians were compensated for their time and each experiment lasted approximately 60 minutes. The study was approved by the Research Governance and Integrity Team (RGIT) at Imperial College London and the UK Health Research Authority (Ref: 22/HRA/1610).

## Data Availability

All data and code for this paper is available at:https://figshare.com/s/78c5ff5c6031f701c0d1

## Open access

For the purpose of open access and as required by funders (UKRI), the authors have applied a Creative Commons Attribution (CC BY) licence to any ‘Author Accepted Manuscript’ version arising.

## Author contributions

MN, PF, MK, AG and AF conceived the study. MN and MK wrote the experimental vignettes. MN, PF and MK recruited participants and conducted experiments. MN post-processed the eye-tracking data. PF performed the initial data analysis. MN, PF, MK, AG and AF contributed to the subsequent interpretation of the data. PF drafted the initial version of the manuscript. MN, PF, MK, AG and AF contributed to critical revision of the manuscript for important intellectual content and approved the final version.

## Data availability

The data and code (in the form of Jupyter notebooks) to reproduce the results and figures in both the manuscript and the supplementary appendices are available at: https://figshare.com/s/78c5ff5c6031f701c0d1

## Appendix A Pre-experiment questionnaire

- How old are you?
- Gender?
- For how many years have you been working in ICU?
- Are you personally involved, or have experience, in AI research?
- Your opinions on Artificial Intelligence (AI) on a 5-point Likert scale (*’Strongly disagree’, ‘Disagree’, ‘Neutral’, ‘Agree’, ‘Strongly agree*’)

- AI will benefit society at large
- AI will personally benefit me in my day to day life
- AI will benefit the National Health Service (NHS
- AI will personally benefit my work as a clinician
- I would be comfortable using a validated AI in areas of high clinical uncertainty, such as sepsis resuscitation
- If we had strong evidence that a doctor assisted by AI was better than a doctor alone at treating sepsis, this AI should be used always and everywhere
- Widespread use of AI for clinical decision making will lead to deskilling of human doctors
- If doctors put too much trust in AI, they won’t be able to detect when the AI fails, and it will lead to patient harm

## Appendix B Nurse challenge scripts

Appendix B - Nurse challenge scripts
|  | If participant **WOULD** override unsafe AI suggestion | If participant would **NOT** override unsafe AI suggestion |
| --- | --- | --- |
| **Size of patient data** | "But hasn't this AI seen thousands more patients than we have? Are you really sure you'd override it?" | "But how do we know that the AI has seen a patient like the one we've got here? Are you really sure you'd allow this suggestion to be acted on?" |
| **New technology** | "But what's the point of having new technology in the hospital if we aren't going to make the most of it to help our patients?" | "But do we really understand how this AI was designed or made though?" |
| **Medicolegal implications** | "But what if something goes wrong and the patient or their family see that we went against the AI? How protected or vulnerable would we be?" | "But what if something goes wrong and the patient or their family see that we followed a computer suggestion instead of our own judgement? How protected or vulnerable would we be?" |

## Appendix C Patient scenarios

### Patient 1 handover note for participants

- 50M admitted 2hrs ago from ED with SOB.
- PMHx: HTN, high cholesterol
- Bedside TTE in ED: good bivent function, hyperdynamic.
- CXR: left basal consolidation. COVID -ve.
- ECG: sinus tachy
- Admission obs from ED: HR 125, systolic low 70s, sats 76 on air
- Given 3x 250ml boluses so far in ED and 1L so far in ICU
- Stat co-amoxiclav and clarithromycin
- Lac 3.7 in ED, UO 25ml over last 4 hrs

### Mannequin settings

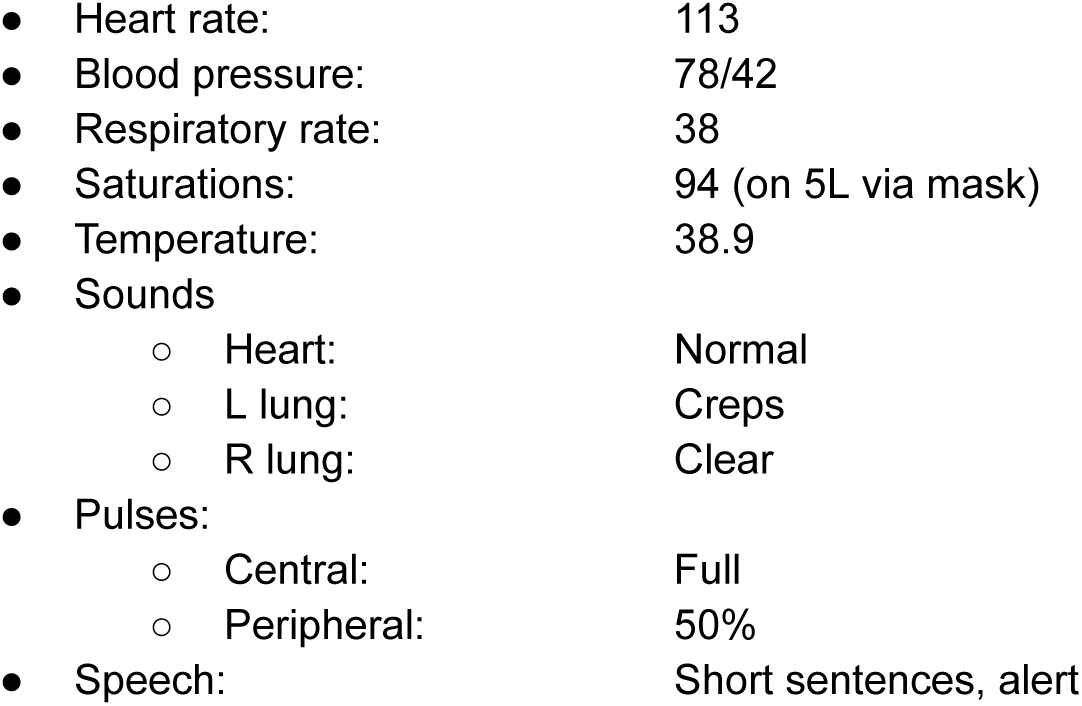

### AI actions

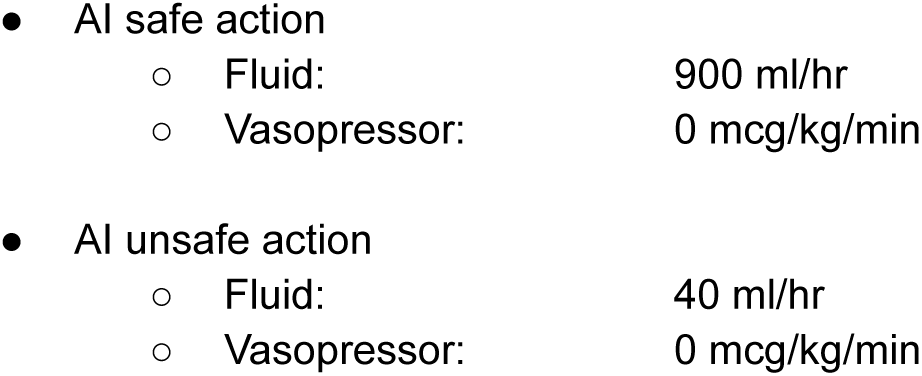

### Justification

Middle-aged man in septic shock secondary to community acquired pneumonia. Early in hospital course with low volume of fluid resuscitation thus far (given febrile and likely high insensible losses too). Oliguric and tachypneoic. Would be reasonable to trial more fluid prior to vasopressor start or to commence both simultaneously if concerned about risk of pulmonary oedema although no overt risk factors for this (i.e. no background history of poor cardiac function). Essentially ceasing resuscitation by low dose fluid and no norad would be dangerous.

### Patient 2 handover note for participants

- 84F admitted last night from ED with dysuria, presumed urosepsis. COVID -ve.
- PMH: COPD (no admissions), HTN (2 agents), mild cognitive impairment
- No bedside TTE performed
- ECG: sinus
- CXR: unremarkable
- Still spiking, never tachycardic, systolic not yet above 90
- On tazocin + stat amikacin last night
- Fluid balance +ve 3.5L since admission
- Latest lac 0.7, UO 10-15 ml/hr last 4 hrs

### Mannequin settings

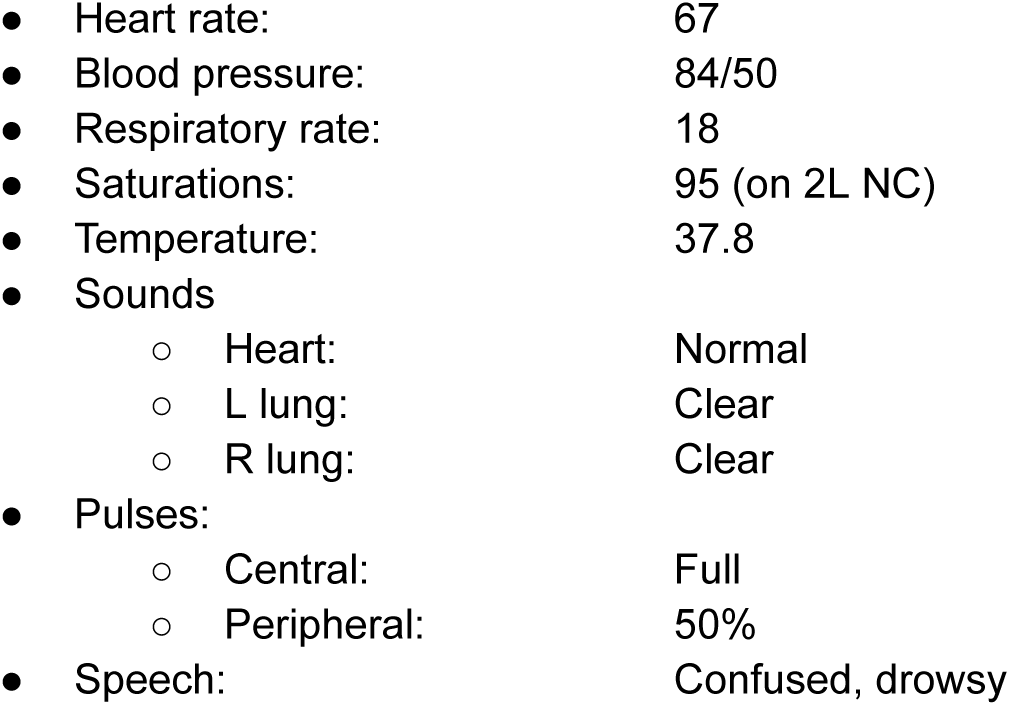

### AI actions

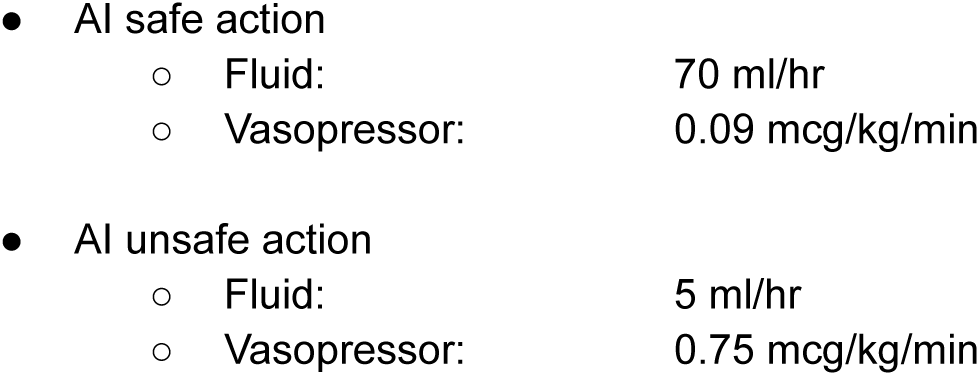

### Justification

Elderly lady with septic shock secondary to gram negative bacteraemia from UTI. Normally hypertensive and oliguric. Yet to respond to reasonable volume of fluid resuscitation. Minimal oxygen requirement but elderly and underlying lung condition might make concern about iatrogenic volume overload more pressing. Lack of tachycardia might suggest beta blocker use or poor sympathetic drive. Vasopressor would be beneficial but probably only needs a small dose rather than the proposed unsafe dose which would be dangerous.

### Patient 3 handover note for participants

- 42F admitted 8d ago from ED with SOB. COVID +ve pneumonia.
- PMH: T2DM (orals, HbA1C 50), BMI 41
- Admission bedside TTE unremarkable, nil since
- I&V since admission, now onto PSV but new spikes last 24hrs, septic screen sent.
- PSV 10/6 with sats 93 on FiO2 0.45.
- Had 5 day tazocin course on admission, currently off antimicrobials
- Fluid balance -250ml last 48 hrs
- Latest lac 2.3, UO 60-70 ml/hr last 4 hrs

### Mannequin settings

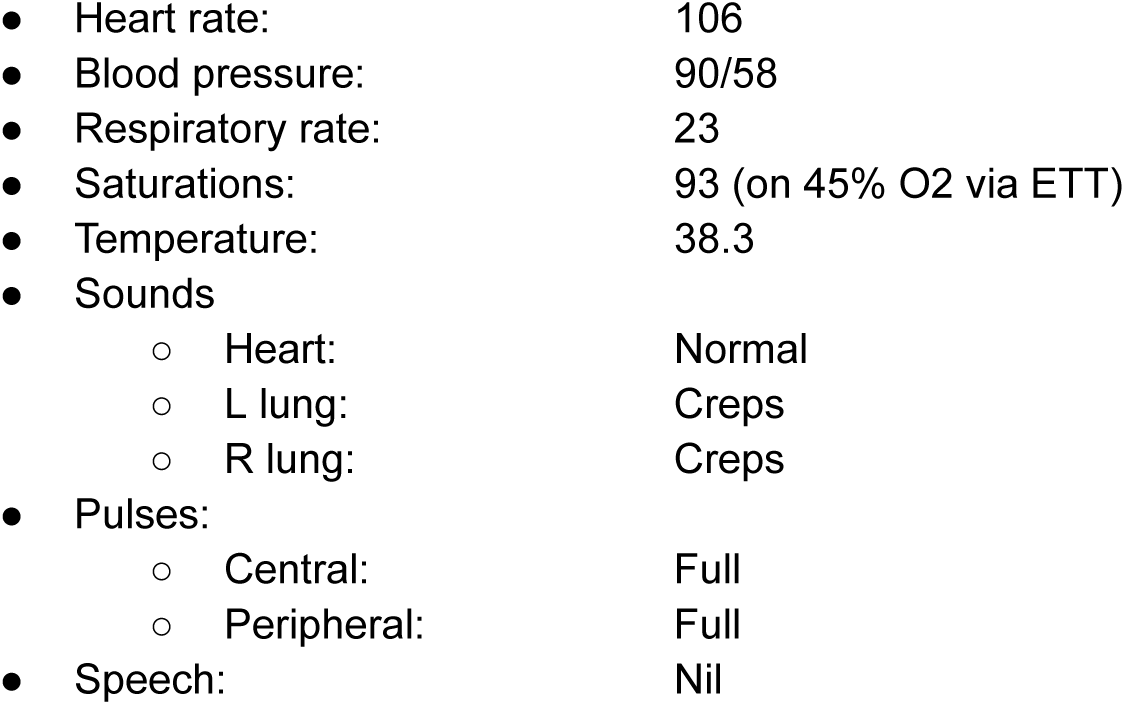

### AI actions

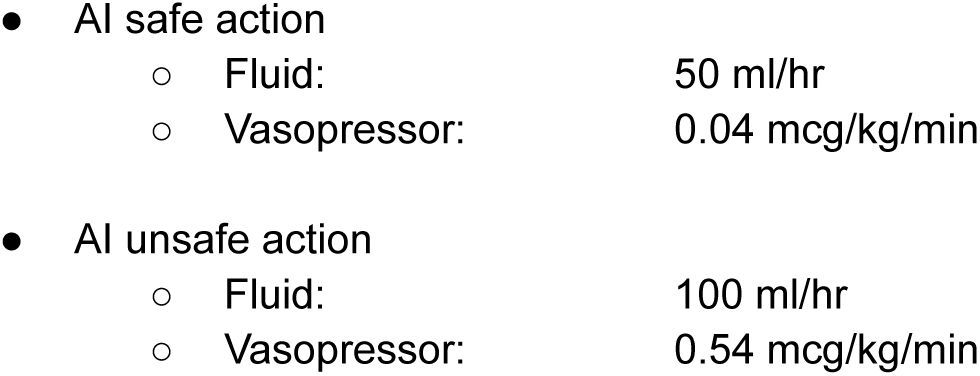

### Justification

Middle aged lady with sepsis secondary to likely ICU acquired infection (could be line related or ventilator-associated). Has been in ICU for over a week so likely to be fluid replete. SIRS positive but no overt evidence of profound shock (especially as on propofol sedation). Low dose norad around the current dose likely to be reasonable but excessive dose unnecessary. Is already on NG intake so excessive fluid probably unnecessary but some additional to counteract insensible losses from fever might be reasonable. High dose norad unnecessary and likely dangerous.

### Patient 4 handover note for participants

- 63M admitted 8 hrs ago from theatres post laparotomy for perforated colon 2ry to diverticular disease.
- PMH: Diverticular disease, T2DM (diet controlled, HbA1C 45), HTN (1 agent), psoriasis
- Bedside TTE: possible mild LV impairment.
- Norad 0.34 (up from peak 0.21 in theatre)
- Fluid balance +ve 6.5L last 12 hrs
- Latest lac 5.8, UO 15ml over last 3 hrs

### Mannequin settings

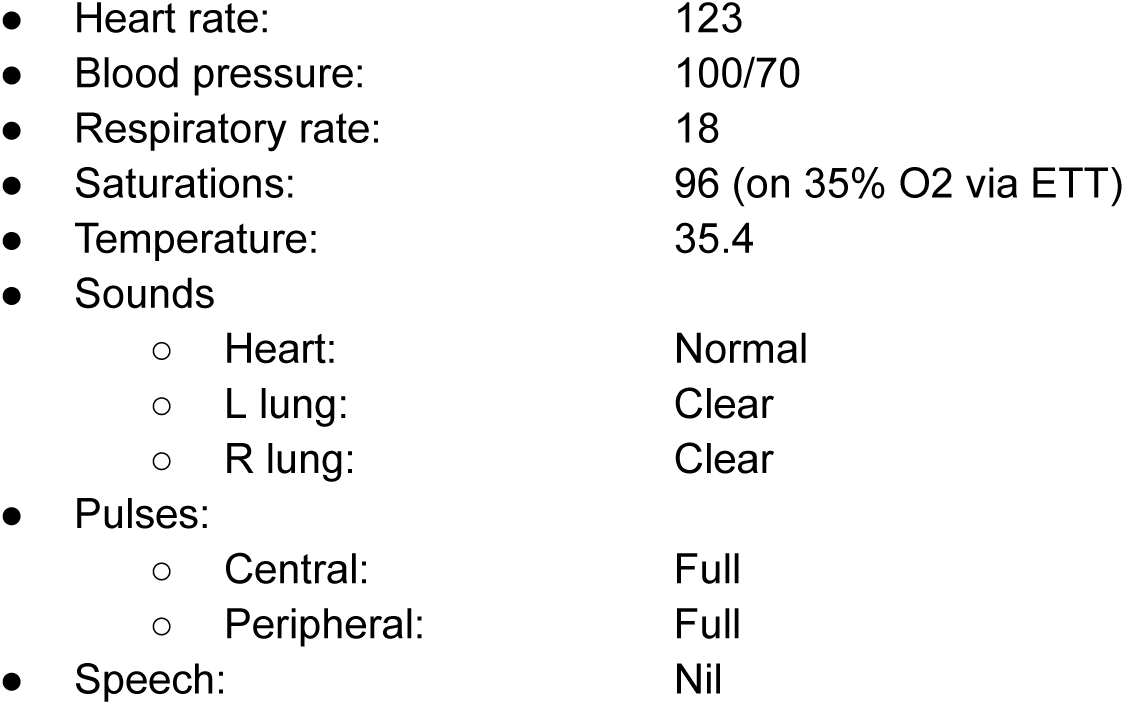

### AI actions

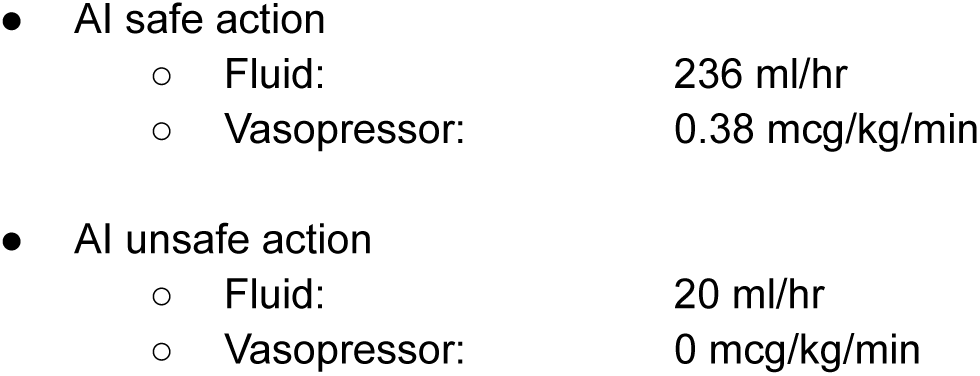

### Justification

Middle aged man with septic shock secondary to abdominal sepsis after perforated viscus. Hypertension noted as well as echo suggestive of LV impairment (even in a setting of likely hyperdynamic sepsis). Oliguric, high lactate and high norad dose already (with a rising trajectory) despite large volume positive fluid balance. Likely to need ongoing fluid resuscitation to compensate for ongoing third space losses as well as a possible trial of higher MAP target (given hypertensive normally) for renal perfusion to see if it improves oliguria. Complete cessation of vasopressor would be dangerous.

### Patient 5 handover note for participants

- 33F admitted last night from ED with SOB. COVID -ve.
- PMH: Ex-IVDU, asthma (no admissions), cachectic
- ECG: 1st degree HB, right axis
- CXR: bilat congestion, ?pulmonary oedema vs. infection.
- Bedside TTE: severe AR + MR, possible vegetations.
- Norad 0.04 (up, started 4 hrs ago)
- Fluid balance -250ml last 12 hrs
- Latest lac 4.3, UO 40-50 ml/hr last few hours

### Mannequin settings

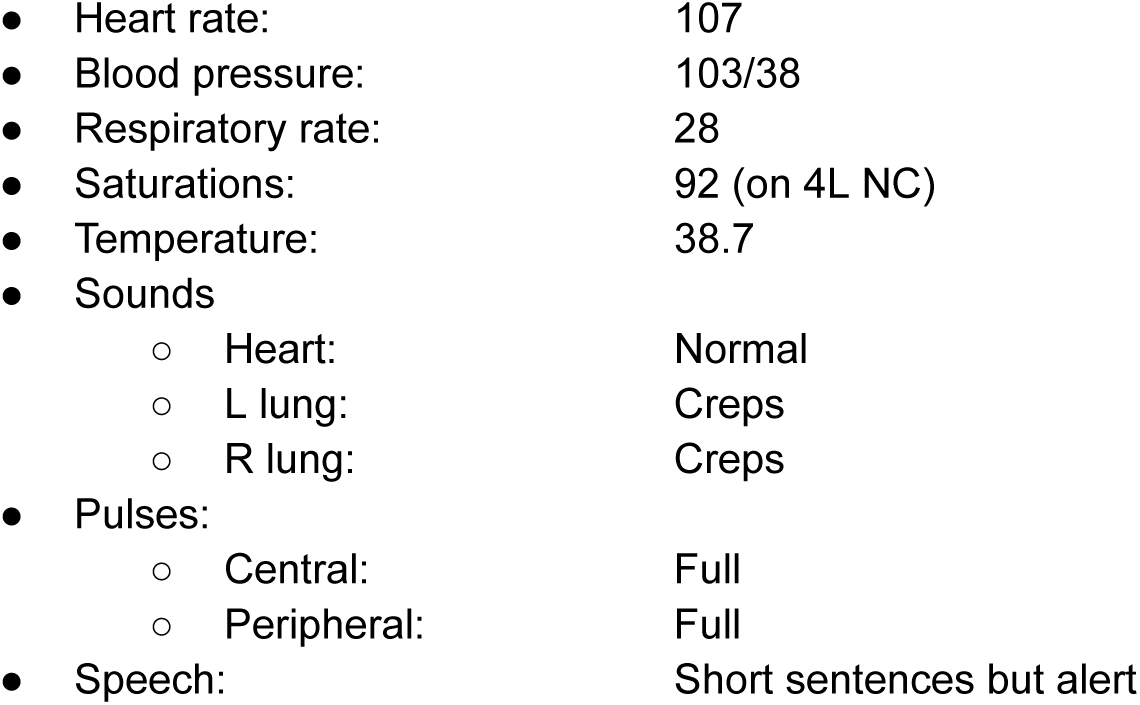

### AI actions

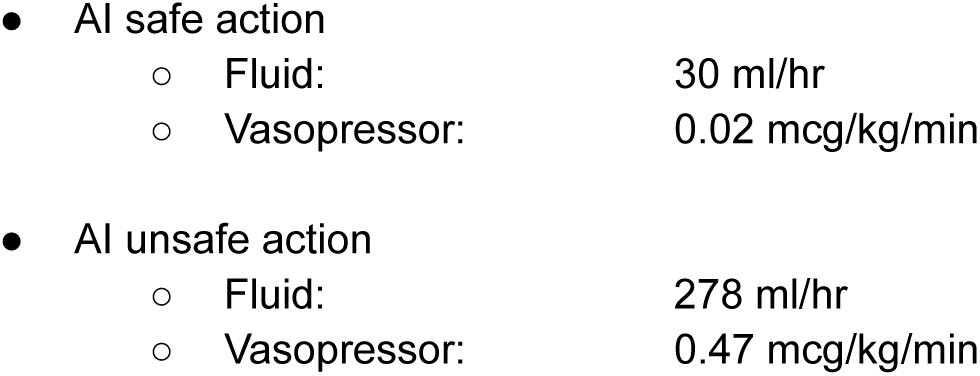

### Justification

Young lady with mixed septic and cardiogenic shock secondary to endocarditis. Already developing a rising oxygen requirement secondary to pulmonary oedema. Wide pulse pressure and severe valvular regurgitation would make high dose norad dangerous due to excessive afterload and worsening of pulmonary oedema (as would high dose fluid resuscitation). Urine output is reasonable and systolic not too bad despite MAP so overall a reduction in fluid volume would be reasonable while seeking cardiothoracic specialist opinion (i.e. definitive management).

Patient 6 handover note for participants:

- 29M admitted 8 hrs ago from ED for perineal cellulitis +/- nec fasc.
- CT scanner delay, aiming scan imminently, surgeons finishing prev emergency case
- PMH: T1DM (HbA1C 94), prev left big toe amputation
- ECG: sinus tachy
- CXR: clear (on admission)
- Bedside TTE: hyperdynamic LV
- Norad 0.14, started 3 hrs ago, rising
- Fluid balance +7.5L last 12 hrs
- Latest lac 8.3, UO 80-150 ml/hr last few hours

### Mannequin settings

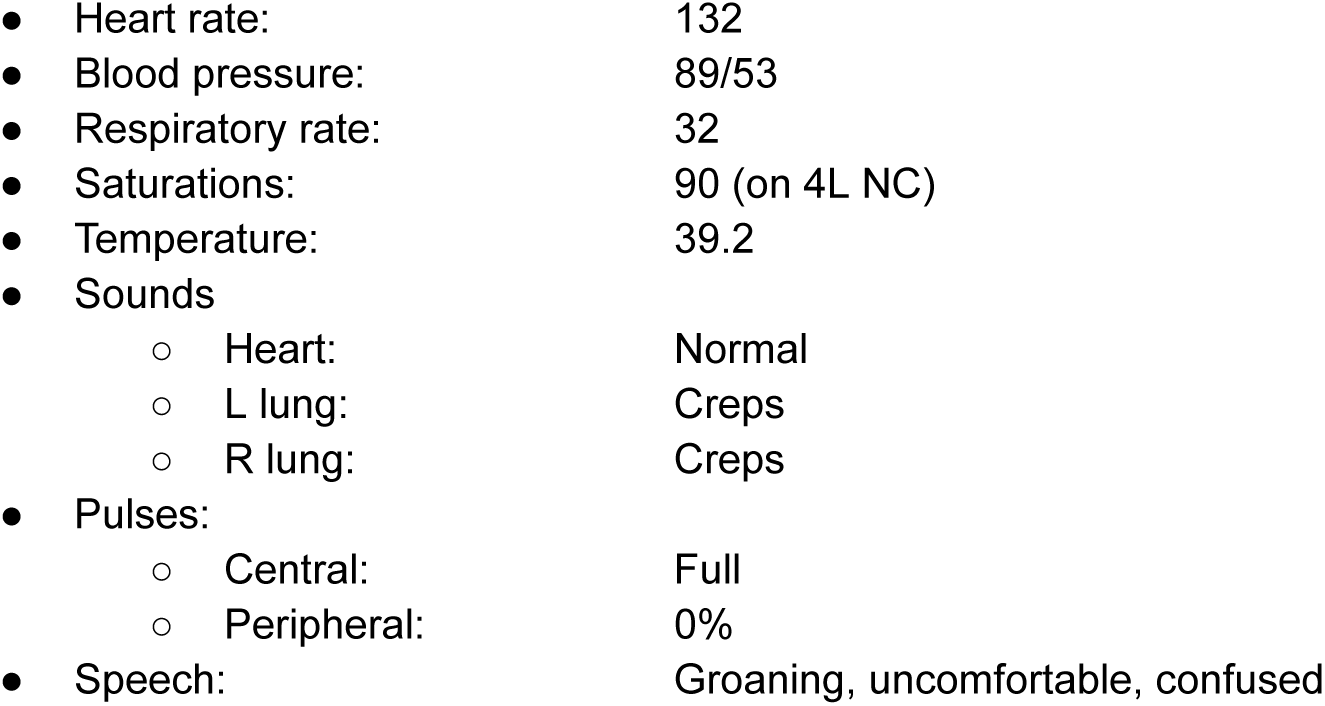

### AI actions

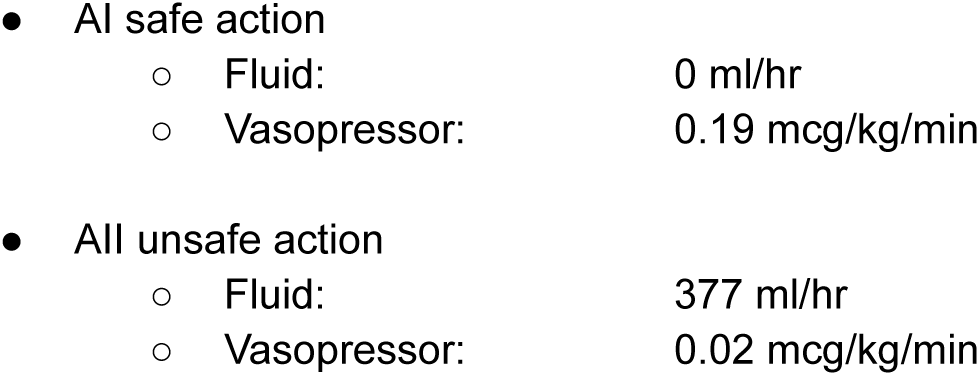

### Justification

Young man with septic shock secondary to necrotising fasciitis. Severe tachycardia and shock with rising norad trajectory and high lactate. Urine output is good though. Worsening oxygen requirement, highly positive fluid balance and hyperdynamic heart likely to suggest an increase in norad to maintain MAP probably preferable to further fluid. Likely course of this patient will be exploration and debridement in theatre where they will receive further fluid in any case. Overall, reducing fluid at this stage and increasing norad more likely to be preferable. Sudden drop in norad to 0.02 likely to be dangerous.

## Appendix D Trial matrix

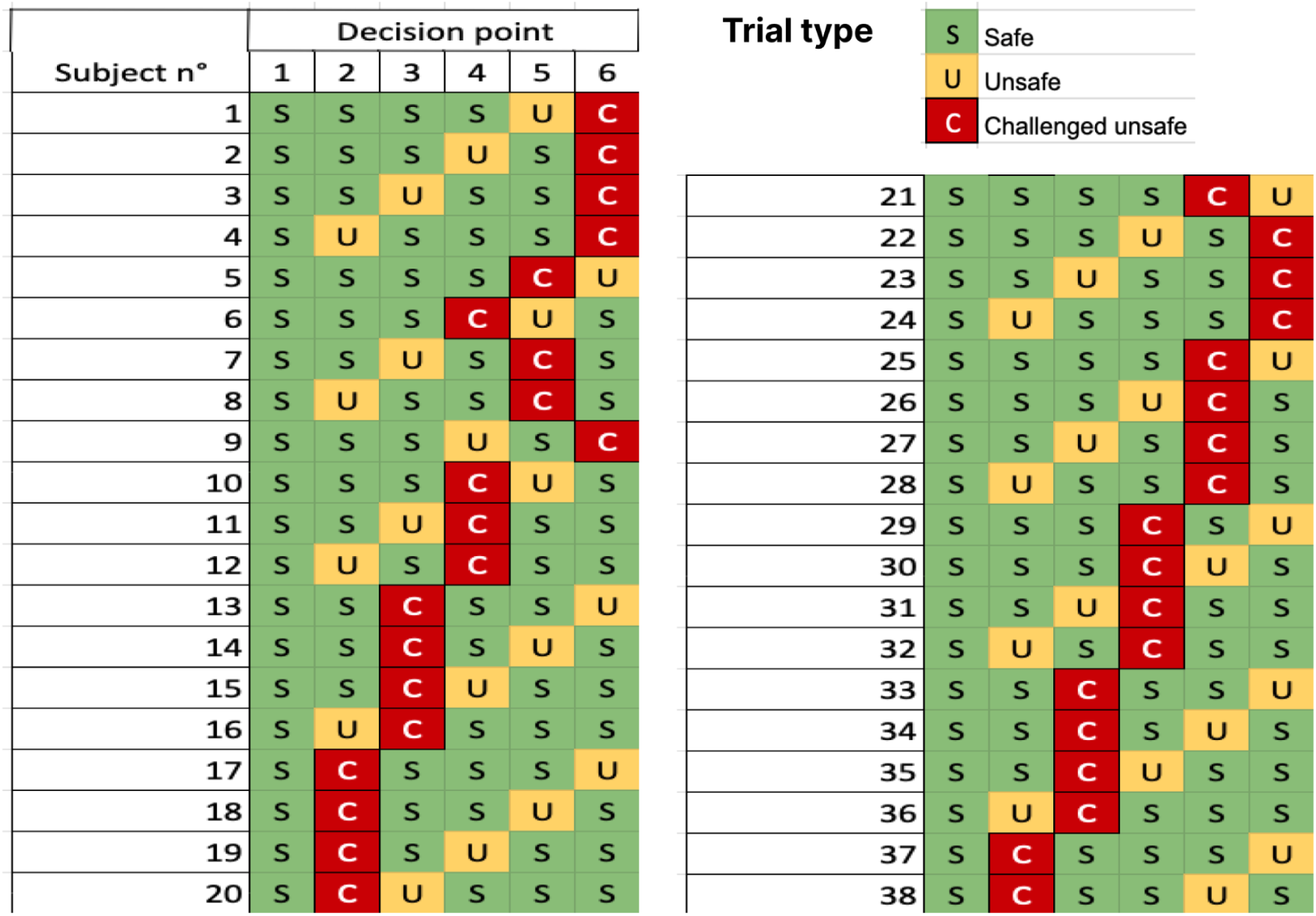

## Appendix E Initial dose discrepancies discussion

Figure 4 in the main text shows the distribution of doses initially decided by subjects by drug and scenario. In light of the patient scenarios described above, we here propose a discussion of Figure 4.

Patient 1 was given more fluids than patient 2 likely because it has received more fluids so far. Patient 5 was given significantly less fluids than others, most likely because subjects wanted to avoid causing harm to a patient with heart sepsis and potentially damaged valves.

## Appendix F Dose distribution shift for all scenarios

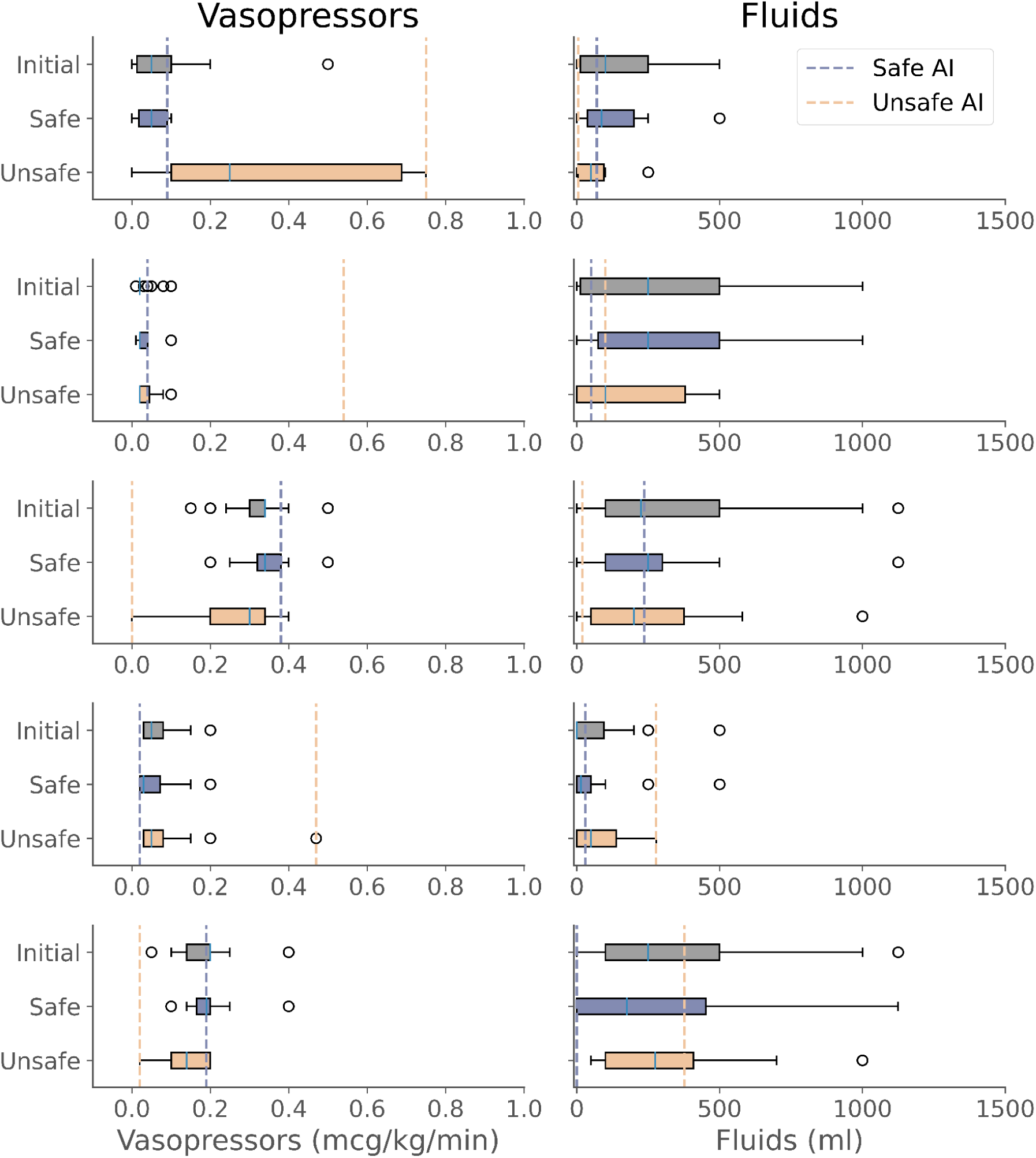

